# An Expert-guided Hierarchical Graph Attention Network for Post-traumatic Stress Disorder Highly-associative Genetic Biomarkers Identification

**DOI:** 10.1101/2023.01.30.23285175

**Authors:** Qi Zhang, Yang Han, Jacqueline CK Lam, Ruiqiao Bai, Illana Gozes, Victor OK Li

## Abstract

Post-traumatic Stress Disorder (PTSD) is a common debilitating mental disorder, that occurs in some individuals following extremely traumatic events. Traditional identification of Genetic Markers (GM) for PTSD is mainly based on a statistical clinical approach by comparing PTSD patients with normal controls. However, these statistical studies present limitations, often generating inconsistent results. Few studies have yet examined thoroughly the role of somatic mutations, PTSD disease pathways and their relationships. Capitalizing on deep learning techniques, we have developed a novel hierarchical graph attention network to identify highly correlational GM (HGMs) of PTSD. The network presents the following novelties: First, both a hierarchical graph structure and a graph attention mechanism have been integrated into a model to develop a graph attention network (GAtN) model. Second, domain-specific knowledge, including somatic mutations, genes, PTSD pathways and their correlations have been incorporated into the graph structures. Third, 12 somatic mutations having high or moderate impacts on proteins or genes have been identified as the potential HGMs for PTSD. Fourth, our study is carefully guided by prominent PTSD literature or clinical experts of the field; any high saliency HGMs generated from our model are further verified by existing PTSD-related authoritative medical journals. Our study illustrates the utility and significance of a hybrid approach, integrating both AI and expert-guided/domain-specific knowledge for thorough identification of biomarkers of PTSD, while building on the nature of convergence and divergence of PTSD pathways. Our expert-guided AI-driven methodology can be extended to other pathological-based HGM identification studies; it will transform the methodology of biomarker identification for different life-threatening diseases to speed up the complex lengthy procedures of new biomarkers identification.

## Introduction

Post-traumatic Stress Disorder (PTSD) is a common and debilitating mental disorder that occurs in some individuals following exposure to extremely traumatic events, such as life-threatening accidents or natural disasters ^1,2^. It leads to symptoms such as re-experiencing (e.g. having trauma-related memories that intrude into what is currently happening), avoidance of stimuli associated with the trauma, negative changes in cognition and mood, and hyperarousal ^2-7^. These symptoms could cause serious and long-lasting problems, including unemployment, marital instability, physical illness, and early mortalities ^3,8-14^. Family, twin and molecular genetic studies have suggested that genetic factors contribute to the development of PTSD ^13-23^. However, despite more than a decade of research efforts, robust Highly-associative Genetic Markers (HGMs) of PTSD remain largely unknown ^4^.

Traditional methods identifying genetic markers related to PTSD are mainly based on statistical analysis and comparisons among PTSD patients and normal controls ^2,4,11,14,17,20-22,24-50^. Two major approaches are candidate gene association studies and Genome-wide Association Studies (GWAS) ^2,4,11,14,17,20-22,24-54^.

In candidate gene association studies, only a few selected genetic markers are involved in the analysis, and the selection is mainly based on existing biological knowledge obtained from prior research on PTSD-related neurobiological processes ^17,20,24-26^. Much research efforts have been made in candidate gene association studies ^27-43^. For instance, the FKBP5 gene, which is an important regulator of the stress system, has been suggested to have single-nucleotide polymorphisms associated with PTSD through interactions with child abuse ^3,27,31^. However, the majority of PTSD HGMs could hardly be identified by candidate gene association studies, since such studies are usually limited to genetic regions where there are prior hypotheses about their roles in the development or maintenance of PTSD, whereas people’s prior understanding on the pathophysiology of PTSD is incomprehensive or even incorrect ^14,17,21^.

In GWAS, the frequencies of hundreds to millions of genetic variants across the entire genome are compared simultaneously between those with and without PTSD ^17,20,21,24,25,44,45^. It is a hypothesis-free approach avoiding the need for prior knowledge of specific candidate genes/variants, and thus capable of identifying unknown mechanisms ^20,24,45^. It has gained momentum in recent years for PTSD-related genetic marker identification ^2,4,11,14,22,24,46-54^. For example, a GWAS conducted on veterans and their intimate partners reported a genome-wide significant association between PTSD and a single-nucleotide polymorphism (rs8042149) located in the RORA gene ^46^. However, given the large number of genetic markers simultaneously involved in the statistical analysis, large sample sizes are required for GWAS to have statistical power ^44,55^. Results of GWAS are also best combined with known or putative PTSD-related biological knowledge, in order to avoid spurious findings caused by sampling bias ^20^.

There are other problems with traditional genetic studies of PTSD. A major issue is the inconsistency among research results when different samples are investigated ^4,21,25,26,41,43,56-59^. Besides, previous PTSD-related studies have mainly focused on germline mutations, with the underlying hypothesis that PTSD-related genetic factors are heritable, while less attention has been paid to somatic mutations ^2,4,11,14,22,27,30-33,37-40,42,43,47-54,56,59^. In 2021, Sragovich et al. have extracted somatic mutations from RNA-seq data, and utilized STRING analysis, which is a method based on protein interaction information, to identify crucial PTSD-related genes with somatic mutations^60^. Eight genes have been identified in the study, including TSC1, FMR1, GSK3B, EZR, TNF, IL1R2, CASP1 and CASP4 ^60^. However, studies in this field are still in the beginning stage.

In recent years, Artificial Intelligent (AI) techniques have been utilized in finding genetic markers associated with diseases, though not on PTSD. For instance, to classify whether a gene is associated with Parkinson’s Disease (PD), a neural network-based ensemble (n-semble) method based on protein features has been put forward, reaching 88.9%, 90.9% and 89.8% for the precision, recall and F score in a five-fold validation, respectively ^61^. Another PD-related gene prediction model named N2A-SVM has also been proposed based on protein interaction information and techniques including Node2vec, the autoencoder and the support vector machine ^62^. Its area under the receiver operating characteristic (ROC) curve reaches 0.7289 for classifying whether a gene is associated with PD in a ten-fold validation ^62^. A similar methodology has also been applied to the disease Multiple Sclerosis (MS), and achieved 70.11% accuracy for classifying whether a gene is associated with MS in a five-fold validation ^63^. Besides, Chang et al. ^64^ has proposed a deep learning method based on a sparse auto-encoder to identify cancer-related genes, by extracting features from protein expression profiles and protein interaction information, and achieves ROC value over 0.8 in predicting cancer-related genes. There are also studies applying AI methods for identifying Alzheimer’s Disease (AD)-related genes, using techniques including autoencoders, stepwise artificial neural networks, convolutional neural networks and conditional generative adversarial networks ^65-68^. Given the lack of ground truth, most of those papers have not reported their exact accuracy for identifying AD-related genes, while some of them have claimed that previous studies/analyses (related to AD/neurodegenerative diseases or gene functions) could support some genes they identified ^65-68^. Besides, in 2021, Li et al. have published a plan on designing an AI-driven causal graph model to identify the HGMs for AD in the future ^69^. Moreover, utilizing data on 83 diseases, a feed-forward neural network has been designed for disease diagnosis based on information of gene expression and disease pathways, and sensitivity analysis has been performed to identify associations between diseases and genes ^70^. There are literature supports for 70% of the top 10 disease-gene associations identified in the study ^70^. Thus, AI techniques seem promising in identifying genetic markers for diseases.

In this paper, we have developed a graph-based deep learning diagnosis model to identify probable HGMs for PTSD, utilizing hierarchical graph structures and graph attention mechanisms ^71,72^. Compared to previous studies, our novelties are listed as follows:

- We have constructed a hierarchical biomedical graph representing different layers of one’s biological system, ranging from somatic mutations, genes, to pathways, while incorporating a variety of domain-specific knowledge during the graph construction process, including the impact level of somatic mutations to proteins (i.e. CADD scores), lengths of genes, and the number of PTSD-related pathways on which each gene locates.
- We have utilized graph attention mechanisms to calculate the weights of complex gene-gene interconnections to complement the domain-specific pathway-based information. We have also used attention mechanisms to capture crucial long genes and genes located in more PTSD-related pathways.
- We have proposed a novel saliency score that calculates PTSD-related risk for somatic mutations utilizing the novel hierarchical graph attention network (H-GAtN). A list of HGMs have been identified based on the saliency score and on the literature.

## Results

### Experimental setting and evaluation

The model was implemented in PyTorch ^73^. The training epoch was 30 and the optimizer was Adam (initial learning rate = 10^−6^, reduced by one-tenth after every 10 epochs). L2 regularization has been applied (weight decay = 10^−7^). The hidden dimensions of the embedding layer, the mutation graph, the gene graph, and the final fully connected layer were 8, 16, 32, and 16, respectively. 80% of the patients and 80% of the normal control subjects were randomly selected as the training set, and the rest were used as the testing set. The percentage of misclassified subjects (the subject would be classified as PTSD patients when the predicted PTSD probability is larger than 0.5, otherwise classified as normal controls) in the testing set was used to evaluate the model’s classification accuracy, and the final error rate could be lowered to 11.8%. The area under the ROC curve was 0.90.

### Top HGMs identification

There were 13566 high/moderate-impact somatic mutations detected in the subjects, which were taken as input features of the model. The studied mutations have high or moderate impacts on the proteins of 4561 genes. After model training, saliency scores of all mutations were calculated. Table 1 shows detailed information about the mutations with the top 20 saliency scores and the gene affected by each mutation. The type of mutations includes insertion-deletion (INDEL), and single-nucleotide variant (SNV).

**Table 1.** Somatic mutations with the top 20 saliency scores

| Ranking<br>(based on<br>Saliency<br>Score) | Chromosome | Position | Type | Reference<br>allele | Alternative<br>allele | Affected gene |
| --- | --- | --- | --- | --- | --- | --- |
| 1 | 8 | 30180593 | INDEL | CAG | C | DCTN6 |
| 2 | 18 | 51176872 | SNV | C | T | MEX3C |
| 3 | 3 | 111606778 | SNV | G | A | CD96 |
| 4 | 11 | 118349865 | INDEL | G | GA | CD3G |
| 5 | 6 | 41198325 | SNV | C | A | TREML2 |
| 6 | 14 | 102050157 | SNV | C | T | DYNC1H1 |
| 7 | 6 | 33008108 | SNV | C | T | HLA-DOA |
| 8 | 7 | 100373707 | INDEL | AT | A | PILRA |
| 9 | 9 | 137039846 | SNV | G | A | NPDC1 |
| 10 | 1 | 158292315 | SNV | G | A | CD1C |
| 11 | 12 | 57741942 | SNV | T | C | AGAP2 |
| 12 | 19 | 51415990 | SNV | G | C | SIGLEC10 |
| 13 | 19 | 51414971 | SNV | C | G | SIGLEC10 |
| 14 | 19 | 54574980 | SNV | A | G | LILRA2 |
| 15 | 7 | 100399296 | SNV | C | T | PILRA |
| 16 | 6 | 32937309 | SNV | G | T | HLA-DMB |
| 17 | 14 | 61549756 | SNV | T | G | PRKCH |
| 18 | 12 | 10315316 | SNV | A | G | KLRD1 |
| 19 | 17 | 27306800 | SNV | T | G | WSB1 |
| 20 | 3 | 10301346 | SNV | C | T | SEC13 |

Among these top 20 mutations, we have identified 12 of them as HGMs based on the related literature. A detailed description of the related literature for these top 20 mutations could be found in Table 3. We selected the HGMs comprehensively considering the closeness of their relationship to PTSD, the number of related research articles, the citations, and the impact factor of the journals. These 12 identified HGMs were further divided into three tiers given the strength of literature support. The identified HGMs and their tiers are listed in Table 2.

**Table 2.**
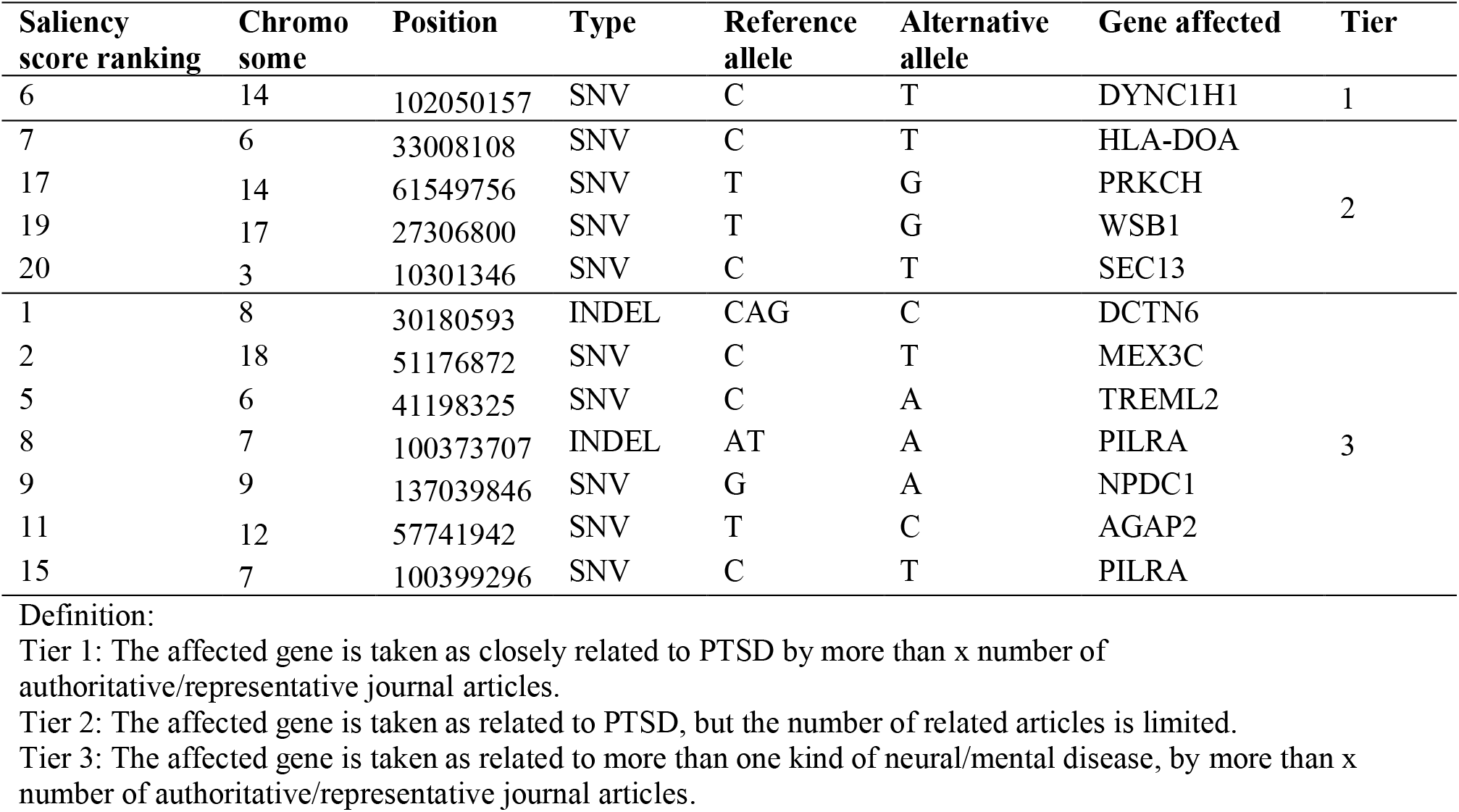
List of Identified HGMs

**Table 3.** A description of the top 20 somatic mutations from representative literature

| Ranking based on saliency score | Affected gene | Related disease | Representative literature |
| --- | --- | --- | --- |
| 1 | DCTN6 | Neural diseases | It was found that DCTN6 deficiency enhances aging in mouse brains<sup>74</sup>. DCTN6 was also identified to have close relation to protein PQBP1, which has been linked to intellectual disability disorders and progressive neurodegenerative diseases<sup>75</sup>. |
| 2 | MEX3C | Mental diseases and neural diseases | MEX3C was identified as one of the risk genes contributing to Neurodegenerative brain diseases<sup>76</sup>. The location of the gene 18q21.2 has been suggested highly correlated with some mental disorders including schizophrenia<sup>77</sup> and depression<sup>78</sup>. |
| 3 | CD96 | Other diseases | CD96 was related to immune system<sup>79</sup>. |
| 4 | CD3G | Mental diseases | CD3G was related to immune system. It was identified to be associated with prenatal depressive symptoms<sup>80</sup>. |
| 5 | TREML2 | Neural diseases | TREML2 was identified to be Alzheimer’s disease risk genes<sup>81</sup>. Missense variant in TREML2 was found to be protective against AD<sup>82</sup>. |
| 6 | DYNC1H1 | PTSD | DYNC1H1 was not only identified as biomarker for diagnosis of PTSD<sup>83</sup>, but also a key target in the treatment of PTSD<sup>84</sup>. |
| 7 | HLA-DOA | PTSD | HLA-DOA was related to immune system. It was identified to be related to PTSD in an article using Transcriptome-wide association studies (TWAS)<sup>85</sup>. |
| 8, 15 | PILRA | Neural diseases | PILRA was identified to be correlated to Alzheimer’s disease by many articles<sup>86,87</sup>. |
| 9 | NPDC1 | Mental diseases and neural diseases | NPDC1 was identified to be correlated to Alzheimer’s disease<sup>88</sup>. It was also identified to be associated with development and prognosis of schizophrenia<sup>89</sup>. |
| 10 | CD1C | Mental diseases | CD1C was related to immune system. It was suggested to have a good diagnostic performance in major depressive disorder<sup>90</sup>. |
| 11 | AGAP2 | Mental diseases | AGAP2 was identified as risk gene related to autism in many research articles<sup>91</sup>. It was also identified as differentially methylated gene related to Alzheimer’s disease<sup>92</sup>. |
| 12, 13 | SIGLEC10 | Other diseases | SIGLEC10 was related to immune system<sup>93</sup>. |
| 14 | LILRA2 | Mental diseases | LILRA2 was identified as differentially expressed transcripts and genes in a study on loneliness<sup>94</sup>. |
| 16 | HLA-DMB | Other diseases | HLA-DMB was mainly related to immune system. It was identified as a gene associated to schizophrenia<sup>95</sup>. |
| 17 | PRKCH | PTSD | PRKCH was identified as a gene significantly upregulated in PTSD cases compared to controls<sup>96</sup>. It was also identified as a gene closely associated with stroke<sup>97</sup>. |
| 18 | KLRD1 | Mental diseases | KLRD1 was identified as possible therapeutic targets of stress-related disorders<sup>98</sup>. |
| 19 | WSB1 | PTSD | WSB1 was identified as a gene significantly upregulated in PTSD cases compared to controls<sup>96</sup>. It was also suggested to be protective towards Parkinson’s disease<sup>99</sup>. |
| 20 | SEC13 | PTSD | SEC13 is part of the complex mTORC1, which is identified as a key to the formation and also the treatment target of PTSD<sup>100,101</sup> and Alzheimer’s disease<sup>102</sup>. |

### HGMs validation

As mentioned in the introduction section, in Sragovich et al.^60^, eight genes with somatic mutations have been suggested to be potentially related to PTSD, including TSC1, FMR1, GSK3B, EZR, TNF, IL1R2, CASP1, and CASP4. Specifically, in the dataset used in our study, there are 12 somatic mutations with high or moderate impacts on those eight genes. We have checked the saliency score ranking of those 12 somatic mutations for reference. Figure 1 shows the saliency scores of all somatic mutations considered in our model, and mutations on the left have higher saliency scores. Vertical red lines correspond to the ranking of the 12 reference somatic mutations. It could be seen that they are distributed across the left-hand side of the figure, which implies the identified risky mutations in Sragovich et al.^60^ also have relatively higher saliency scores calculated by our model. Hence, to a certain extent, our saliency score supports the potential importance of genes identified in Sragovich et al.^60^. The 12 reference mutations are not the ones with top saliency scores in our study. One possible reason is that Sragovich et al.^60^ concentrated on the high-impact mutations while our study focused on both high-impact and moderate-impact mutations.

**Figure 1.**
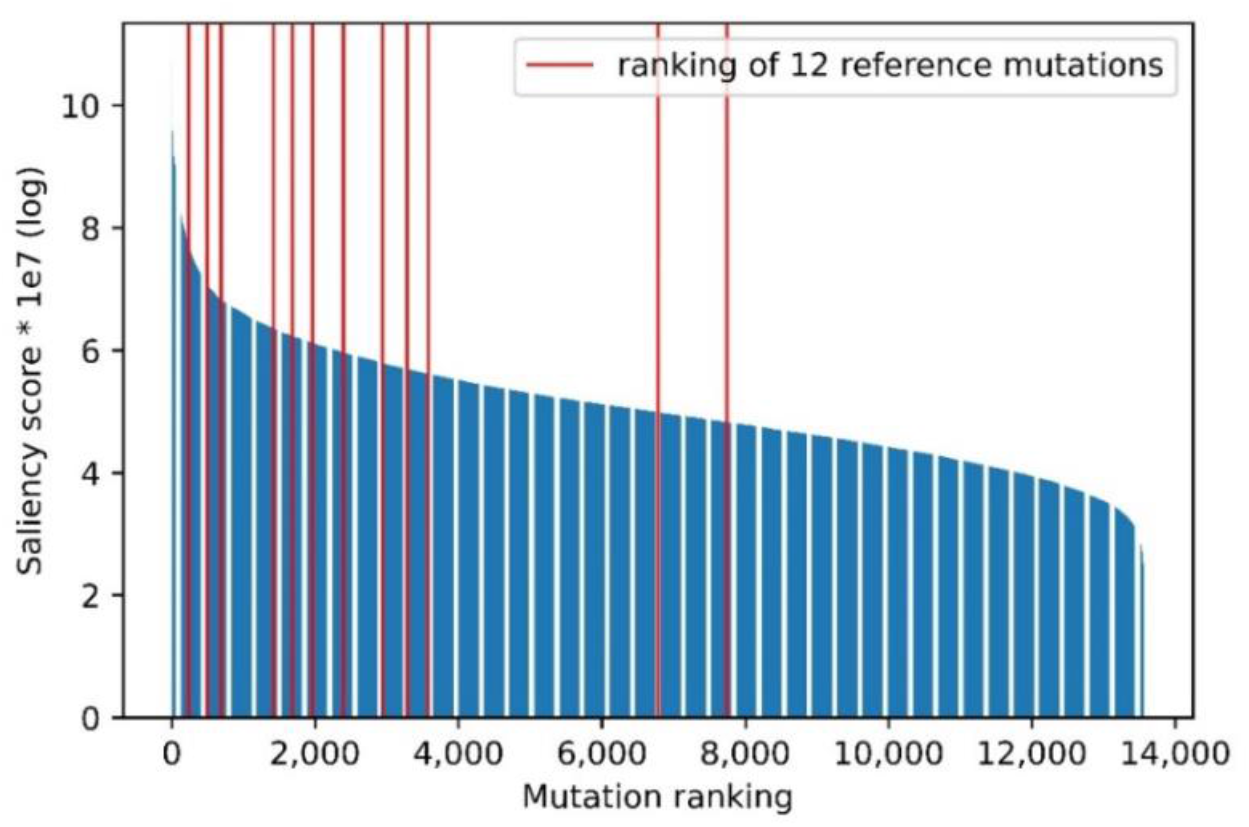
Ranking based on mutation saliency score

To further validate the model’s capability of identifying potential HGMs, we investigated the genes affected by the top 20 HGMs identified by our model, specifically their presence in research related to PTSD or other mental/neural diseases. The supportive literature is summarized in Table 3. It could be seen that most of the genes affected by the top-ranking mutations have also been identified as biomarkers for mental diseases or neural diseases by other researchers. Among them, the gene DYNC1H1 has been recognized by many researchers as not only a potential biomarker but also a key target in the treatment of PTSD. The supportive literature further validates our model’s capability of identifying potential HGMs.

### Network-learned edges

The graph attention convolutional network in our model could learn a set of edge parameters from the training data, which represents the network’s belief in the connection strength between gene pairs. In Fig. 2, we visualized the connectivity of gene node DYNC1H1, which is affected by the top identified HGM. The first-order neighbors are the gene nodes connected to DYNC1H1 with the largest edge parameters. The orange-colored nodes are the overlap of these first-order neighbours and the genes affected by mutations of the top 20 saliency scores. Since the gene DYNC1H1 has been implied to be correlated to PTSD by a sufficient number of research articles, these network-learned neighboring genes could be suggestive candidates for potential PTSD-related pathway studies.

**Figure 2.**
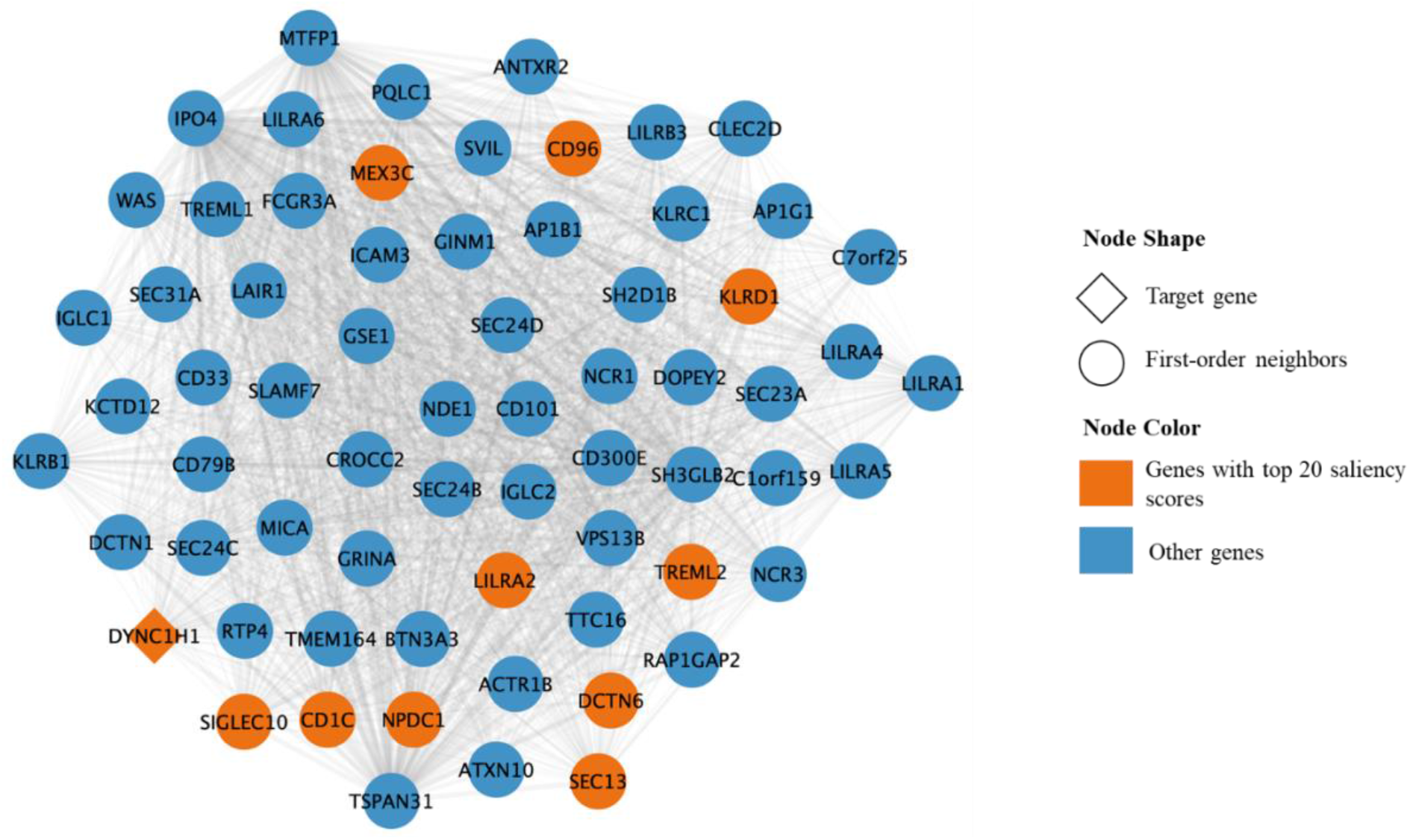
Visualization o network-learned graph edges for gene DYNC1H1

## Discussion

A novel graph-based deep learning diagnosis model has been developed to identify probable HGMs for PTSD. Hierarchical graph structures and graph attention mechanisms have been utilized in the model to incorporate a variety of domain knowledge on somatic mutations, genes, pathways, and their correlations ^71,72^.

Our model has identified 12 mutations as potential HGMs for PTSD, and there are 11 genes whose corresponding proteins are affected by these 12 mutations at high or moderate impact levels. Among the identified high-risk genes, DYNC1H1 was also identified in previous research as not only a biomarker but also a key treatment target for PTSD. For other identified genes, literature support could also be found proving their correlation to PTSD or mental/neural diseases. The learned edge parameters of our graph attention network also provide suggestive candidates for PTSD-related pathway discovery.

It is worth noting that 5 out of the 11 high-risk genes (TREML2, PILRA, NPDC1, AGAP2, SEC13) identified to be related to PTSD by our model were suggested to be closely related to Alzheimer’s Disease in previous research. This implies that there may be underlying connections between these two diseases. Besides, two of our identified genes were suggested to be correlated to depressive symptoms (CD3G, CD1C) and two of them were suggested to be correlated to schizophrenia (NPDC1, HLA-DMB). These findings may provide new insights for future research on not only biomarker identification, but also potential treatment studies like drug repurposing.

Last but not the least, a notable proportion of the top 20 mutations affect genes that are mainly related to the immune system (CD96, CD3G, CD1C, HLA-DOA, HLA-DMB, SIGLEC10). As shown in Table 3, HLA-DMB and HLA-DOA were identified to be associated with schizophrenia^95^. In that study, the authors suggested that their regression analysis supported disease mechanisms that involve the activity of immunity-related pathways in the brain. The similar findings in this study could further demonstrate the importance of the role that immune system plays in the PTSD disease mechanism.

In the future, research efforts are encouraged on improving the HGM identification procedure. A limitation of the current study is the small number of subjects, which might lead to overfitting in the model training. In the future, larger PTSD somatic mutation datasets are in need. In addition, few-shot learning techniques that are tailor-made for HGM identification are suggested to be developed, so as to fundamentally reduce the burden in data collection. Moreover, future studies might also consider the incorporation of quantitative PTSD phenotype measurement, additional demographic information and environmental factors into the model, such as the severity and duration of subjects’ trauma exposure ^50,103,104^.

## Method

### Data Collection and Pre-processing

Two types of data have been collected in this study, including sample data and knowledge-based data. The sample data contained genetic information and demographic information of the studied samples, and were used as the network input. The knowledge-based data contained the biomedical information of mankind, including pathway data and CADD scores, and were used in the construction of the neural network. Figure 3 illustrates our data extraction procedure. Details are specified in the following sections:

**Figure 3.**
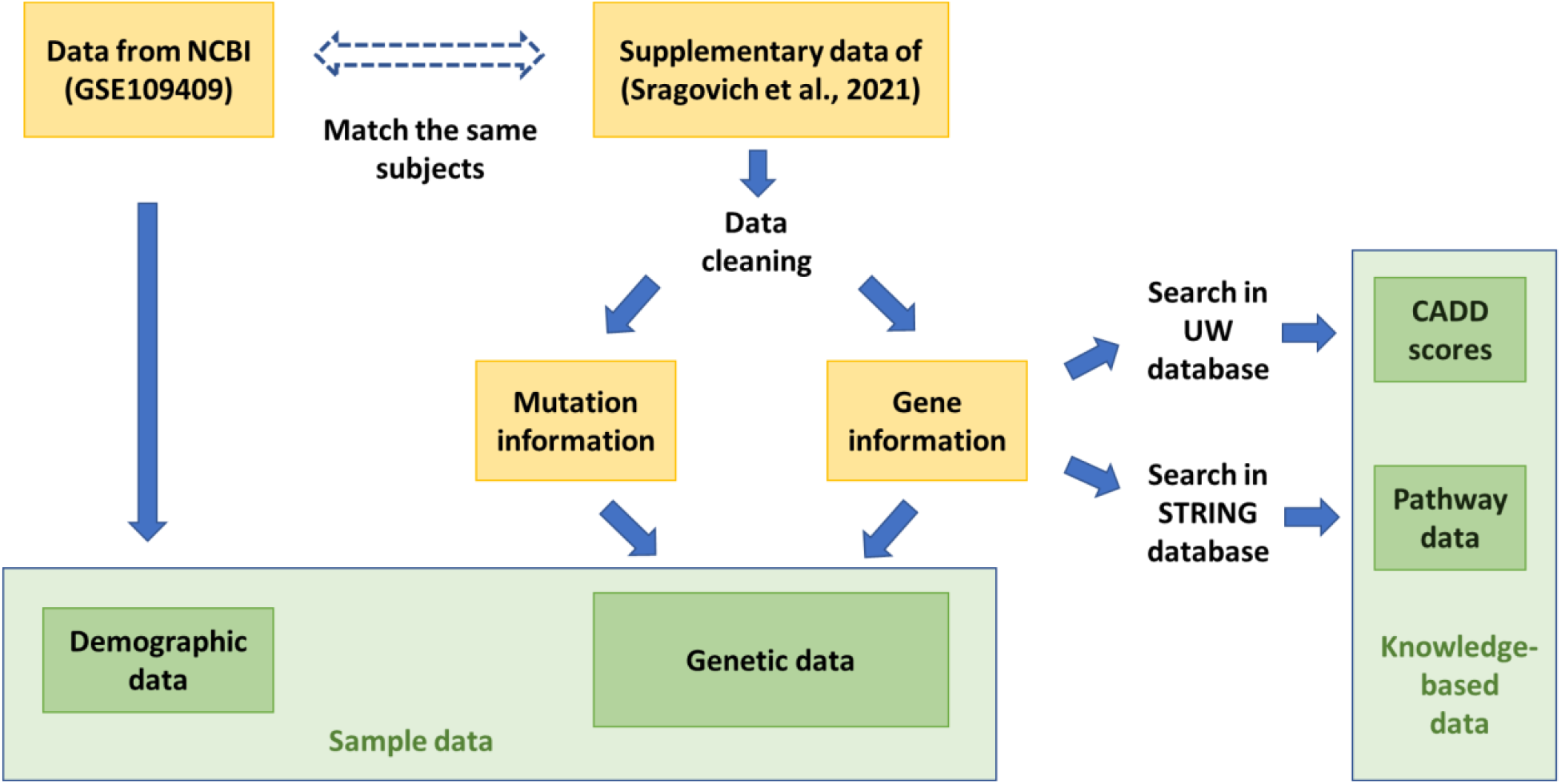
Data extraction procedures: National center for biotechnology information (NCBI), Gene expression omnibus series (GSE), Search tool for retrieval of interacting genes/proteins (STRING), On-line CADD scoring system of Washington University (UW) ^105-108^

### Genetic data

The genetic data were obtained from the supplementary table of a study conducted by Sragovich et al., which extracted somatic mutations from blood samples of 85 Canadian infantry soldiers, including 27 PTSD patients and 58 normal controls ^60^. Specifically, the dataset contains information on somatic Single-nucleotide Variants (SNVs) and Insertions/Deletions (INDELs) with high/moderate impacts on proteins, and information on genes of the proteins they influence. The SNVs and INDELs were mapped to the GRCh38 human reference genome ^60^. Some mismatched columns in the table have been detected and corrected in the data cleaning. Synonyms referring to the same gene in the dataset have been replaced by the latest gene symbol among them using the Ensembl website and MyGene^109,110^. Sragovich et al. ^60^ was based on RNA-seq data of subjects, and RNA-seq read frequencies of SNVs/INDELs have also been extracted from its supplementary table. The SNVs/INDELs without read frequency information in partial subjects (taking up around 1% of all SNVs and INDELs in all subjects) were taken as non-existing mutations in corresponding subjects. Moreover, to account for the fact that somatic mutations are more likely to be implicated in long genes, the lengths of genes were obtained from the Ensembl website and MyGene^109,110^.

### Pathway data

The pathway information was obtained from the STRING database by setting all genes with somatic mutations in the dataset of Sragovich et al.^60^ as inputs, and the STRING database returned pathways that the input genes are enriched in ^105,111^. Specifically, pathways or biological processes from three databases, including the Biological Process (Gene Ontology) database, the KEGG pathway database and the Reactome pathway database, have been extracted ^112-114^. Then, PTSD-related pathways were selected from them by a domain expert to generate the final PTSD-related pathway dataset ^115^.

### CADD scores

The CADD (Combined Annotation Dependent Depletion) score tool^116^ is a logistic regression model for the calculation of variant impact, and it has been utilized to identify genetic markers in previous studies^117^. The CADD tool requires the following variant information: CHROM, POS, REF, and ALT. For the somatic mutations with high/moderate impacts on proteins, we obtained their Phred-scaled CADD scores from the online calculation system provided by Washington University^108^.

### Demographic data

For each subject, we obtained the corresponding demographic information, including age group (six age groups, including 18-24, 25-30, 31-36, 37-42, 43-50, 50-61 years old) and gender, from the original dataset used in Sragovich, et al ^60^ (i.e. NCBI accession number: GSE109409 ^107,118^).

### Ethics approval and consent to participate

Datasets of ^60,107,118^ used in this study were obtained following the research protocol accepted by the Human Research Ethics Committee (HREC) of Defense Research and Development Canada (DRDC) - Protocol 2017-019, and informed consent was obtained from all participants ^60,107,118^.

### Methodology

This study proposed a novel hierarchical graph attention network (H-GAtN) to identify probable HGMs for PTSD. The proposed methodology consists of four steps. Firstly, a hierarchical biomedical graph was constructed utilizing the genetic and pathway data we collected, with nodes corresponding to genes/mutations and edges corresponding to their biomedical connection. Secondly, we trained a hierarchical graph attention network to learn a high-level graph representation of the constructed biomedical graph. Thirdly, the learned high-level feature from the proposed graph neural network was combined with other demographic features, including age and gender. The concatenated features were fed into a deep feedforward neural network for the final prediction of PTSD probability. Finally, we calculated the saliency score for each somatic mutation and obtained the top 20 probable candidates for HGMs.

#### Knowledge-based hierarchical graph construction: mutations, genes, and pathways

A graph as a non-linear data structure can represent the interaction of an arbitrary number of nodes with arbitrary connectivity status, and is thus widely used to model complex real-life scenarios including social networks, traffic forecasting, etc. Previously, graphical convolutional neural networks have also been proven successful in dealing with molecular interaction ^119^ and mutation-related disease prediction ^120,121^. Therefore, we adopted a graph data structure in our PTSD diagnosis scenario, and a biomedical graph was constructed to model the PTSD-related genes and mutations, making it possible for a deep neural network to learn the correlations and interactions of these biomedical concepts.

Specifically, we constructed a hierarchical biomedical graph capturing (1) the interactions of somatic mutations, (2) the interactions of genes, and (3) the hierarchical linkages between somatic mutations and genes (see Figure 4). The first hierarchy of the biomedical graph was a subgraph with each node representing a mutation associated with PTSD. The edges in this subgraph represented the mutation-mutation interactions, and all somatic mutations located on the same gene were linked to each other by undirected edges. The second hierarchy of the graph was a subgraph with each node representing a gene associated with PTSD. The edges in this subgraph represented the gene-gene interactions, and were constructed according to the PTSD-related pathways. All genes involved in the same pathway were linked to each other by undirected edges. The two subgraphs were connected by mutation-gene edges. Each mutation-gene edge connected a somatic mutation to the gene on which the mutation has a high/moderate impact. The weights for the mutation-mutation edges and the gene-gene edges in the subgraph were uniformly set to 1. The weights for the mutation-gene edges were assigned with the corresponding CADD score, and were normalized to the range from 0 to 1.

**Figure 4.**
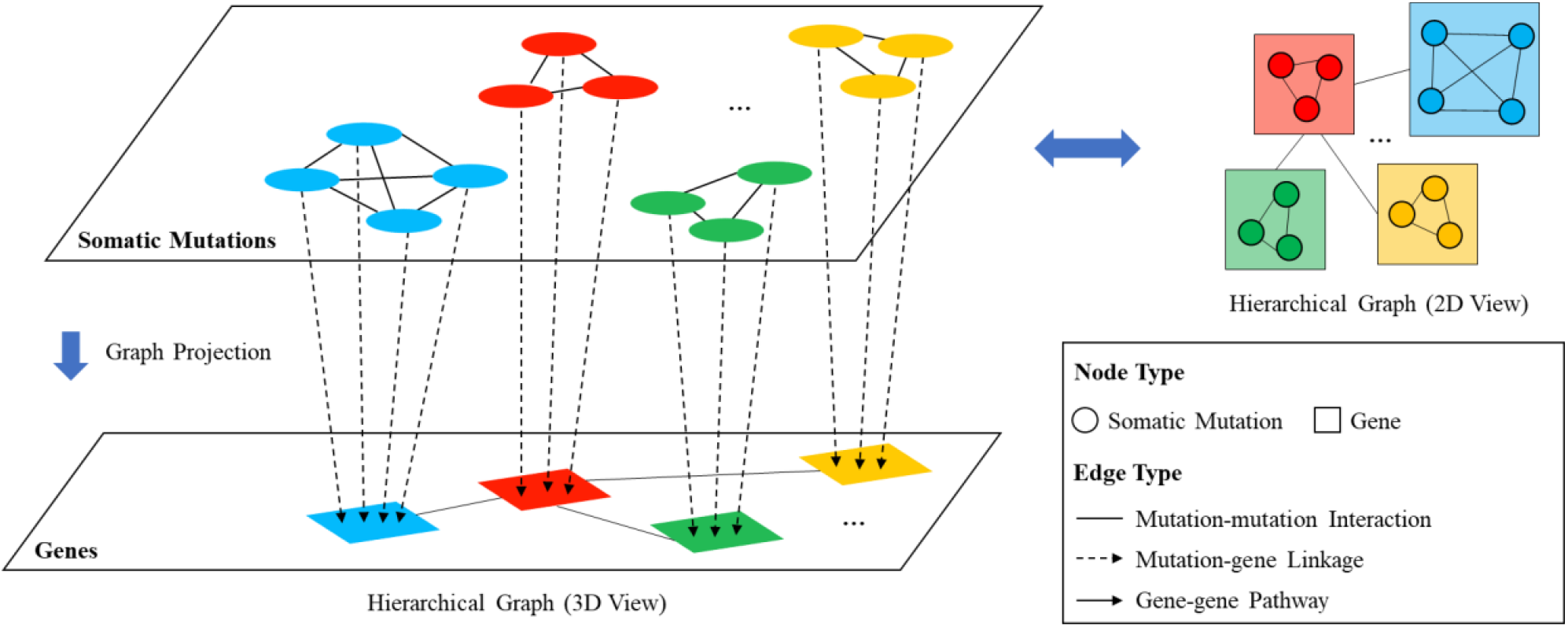
Hierarchical graph construction and projection

#### GNN model structure

After constructing the biomedical graph using domain-specific knowledge, we trained a hierarchical graph attention network to learn a high-level graph representation of the constructed biomedical graph (see Figure 5). The input of the network was the mutation frequency of one subject, and the output was a real number between 0 and 1 indicating the probability of the subject having PTSD.

**Figure 5.**
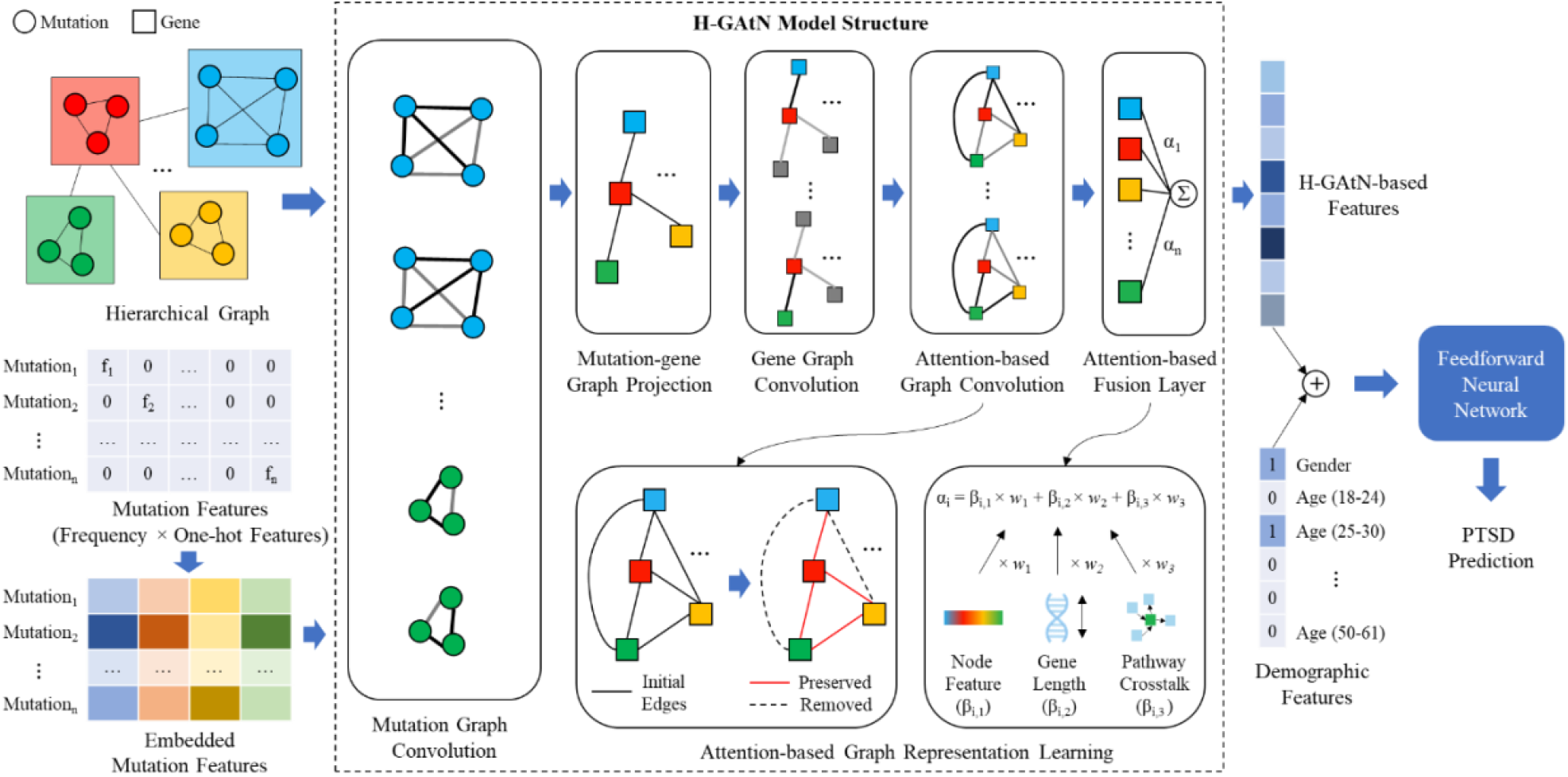
H-GAtN model structure

Each somatic mutation was represented by a one-hot feature vector multiplied by its frequency. The one-hot representations were first embedded into a low-dimensional dense space by an embedding layer. The embedded mutation feature vectors were then fed to the first graph convolutional layer, which conducted graph convolution based on the mutation subgraph. The resulting mutation node features were fed to a graph projection layer, which adjusted the node number and dimension of the node features. Then the projected node features were fed into the second graph convolutional layer, which conducted graph convolution based on the gene subgraph. To better address the complex gene-gene interconnections that are yet to be captured by domain knowledge, a graph attention convolutional layer was incorporated as the third graph convolutional layer. After the hierarchical graph learning, an attention-based fusion layer was incorporated on top of the graph neural network to combine the gene node features and generate a supreme node with its feature vector reflecting the overall information for the subject.

The feature vector learned by our H-GAtN model was then concatenated with the demographic features of the subject. The concatenated vector was fed into a feedforward neural network which generated the final prediction of a real-number probability.

#### Graph projection layer

The graph projection layer was an important structure in our H-GAtN model, designed as a bridge chaining the two graph convolutional layers applied on two subgraphs. The input of the layer was the learned mutation node features ***U***(*n*×*j*), and the layer projected the input into gene node features ***V***(*m*×*i*). n and j are the number of mutation nodes and the dimension of node features in the mutation subgraph. n and i are the number of gene nodes and the dimension of node features in the gene subgraph. Equation (1) illustrates the projection operation. ***E***(*m*×*n*) was a matrix composed of the mutation-gene edges, and left-multiplying ***U*** by ***E*** was equivalent to calculating a weighted sum for mutation nodes connected to the same gene. The mutations with higher CADD scores would have higher weights during the process. *W*(*j*×*i*) was a set of learnable network parameters that transferred the dimension of the resulting node features.

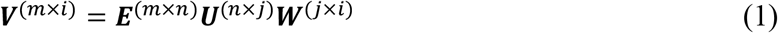

#### Graph attention layer and attention-based fusion layer

Different types of graph convolutional layers were used at different hierarchies of the graph, adapting to the specific characteristics of the data. In the first hierarchy, the vanilla version of graph convolutional layer was applied to process the mutation nodes with known connections. In the second hierarchy, an attention-based graph convolutional layer was also incorporated in addition to the vanilla version of graph convolutional layer. Adopting the GAT layer proposed in Veličković et al. ^72^, the structure of the graph attention convolutional layer is shown in Figure 5. The layer did not require prior knowledge of the pathway information, and all the edges were uniformly initialized to 1. During the training process, the model iteratively optimized its parameters including the edge weights. After training the model on the training data for some epochs, the model preserved the edges that were useful, and removed those useless edges by assigning them a small weight. The graph attention convolutional layer was capable of discovering the pathways that are currently unknown. Stacking it with the vanilla version of the graph convolutional layer, we obtained a network that combined the advantages of the domain-knowledge-based approach and the data-driven approach.

The attention-based fusion layer utilized domain-specific knowledge to merge all the gene nodes into one single supreme node. Our attention mechanism included feature-based attention, gene-length-based attention, and pathway-based attention. Different components of the attention score captured the importance of each gene in different aspects. Equations (2-3) illustrate how the final attention score for each gene node was calculated. *α*_*n*1_ was the feature-based attention score, which measured the similarity of each node feature to a set of learned parameters. *α*_*n*2_ was the gene-length-based attention score for the n_th_ gene, and was determined by the total number of nucleotides within that gene. *α*_*n*3_ was the pathway-based attention score of the n_th_ gene, and was determined by the number of pathways which included that gene.

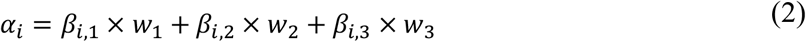

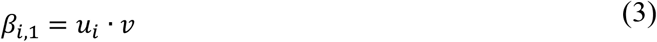

where

*α*_*i*_ is the final attention score of the i_th_ gene,

*α*_*i*1_, *α*_*i*2_, and *α*_*i*3_ are three attention scores of the i_th_ gene calculated in different ways,

*w*_1_, *w*_2_, and *w*_3_ are trainable scalar variables,

*u*_*i*_ is the node vector of the n_th_ gene,

*v* is a trainable vector variable.

#### Demographic features

The learned high-level features from the proposed H-GAtN model were combined with other demographic features, including age and gender. The categorical demographic data were first processed into dummy variables, resulting in a 0/1 vector with 6 components (one for gender information and five for age group information). The demographic features were first rescaled to the same magnitude as the learned H-GAtN-based features, and then concatenated to the learned features. The concatenated vector was fed into a deep feedforward neural network for the final prediction. Ablation study proved that adding the demographic information improved the model accuracy of identifying PTSD patients, reducing the error rate from 17.6% to 11.8%.

#### Saliency analysis

We conducted a saliency analysis to better understand how each mutation could affect the prediction of the final output, thus revealing the importance of each mutation in causing the disease. Previous work in computer vision ^122^ has shown that the gradients with respect to the input values could reflect how much each input feature contributes to the output value. The predicted output (in our case whether the subject has PTSD) of a single subject could be approximated by a linear expression, shown in Equation (4). The magnitude of each dimension of the gradient indicated the relative sensitiveness of that particular input feature. After averaging the gradient on the whole dataset, we defined the saliency score using Equation (5), where *D* is the whole dataset, including training and testing data. After training the model, we calculated the saliency score for each input somatic mutation, and obtained the relative importance of each mutation.

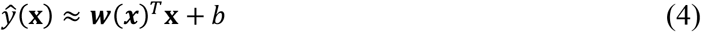

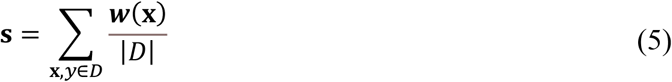

## Data Availability

All data produced in the present study are available upon reasonable request to the authors.

## Data Availability

Partial genetic data supporting this study is available at the supplementary data of Sragovich et al. ^60^. The demographic data used in this article is available at GEO with the accession number: GSE109409. Other datasets generated in this study will be made available upon request to the corresponding authors.

## Code Availability

The code for this study will be made available upon request to the corresponding authors.

## Acknowledgements

This research is supported in part by the US National Academy of Medicine Healthy Longevity Catalyst Award (Hong Kong), 2021 and 2022.

## Author contributions

J.L. and V.L. put forward the research question, the initial methodology, contributed to the design and modification of the model architecture, revised the manuscript, and acquired research funding. Q.Z., Y.H., and R.B. further developed the neural network model for biomarker identification, based on the detailed proposal put forward by V.L. and J.L. R.B. and Y.H. collected and processed the input data. Q.Z. implemented the model and R.B. revised an intermediate version of the codes. Q.Z., R.B., and H.Y. wrote the first draft., I.G. provided the PTSD dataset and information about the PTSD-related pathways. All authors reviewed the manuscript.

## Competing interests

The authors declare no competing interests.

## Notes

### Competing Interest Statement

The authors have declared no competing interest.

### Author Declarations

The study used gene expression data openly available at Gene Expression Omnibus (https://www.ncbi.nlm.nih.gov/geo/) with the accession number: GSE109409.

